# A Pathway-Based Machine Learning Approach Identifies Region-Specific Markers and Patterns in Alzheimer’s Disease Patients Based on Spatial and Severity Metadata

**DOI:** 10.64898/2025.12.01.25341417

**Authors:** Sydney Daynes, Brett E. Pickett

**Affiliations:** Department of Microbiology and Molecular Biology; Brigham Young University; Provo, Utah, United States; 84602

**Keywords:** Alzheimer’s Disease, Machine learning, Signaling pathways, Disease Severity

## Abstract

Alzheimer’s disease (AD) exhibits profound spatial heterogeneity in its molecular and pathological features, yet the basis of this regional selectivity remains obscure. Here, we analyze transcriptomic profiles from publicly available bulk RNA-sequencing datasets collected from two cortical regions implicated in AD vulnerability: the insular cortex and Brodmann area 32 (BA32) of the anterior cingulate cortex. Following rigorous covariate correction and region-specific differential expression analysis, we observe extensive transcriptional disruption in the insula, comprising 4,883 significantly dysregulated genes, while BA32 exhibits no differentially expressed genes at the same statistical threshold. Pathway-level modeling using donor-aware, nested elastic-net logistic regression accurately classified AD versus control cortical samples (pooled BA32 and insula; mean AUROC = 0.813 ± 0.127; AP = 0.938 ± 0.040). SHAP-based interpretability identified a concise set of biologically coherent pathways underlying predictive performance. Cross-regional generalization revealed partial transferability of insular signatures to BA32, indicating shared yet muted molecular changes in the latter. Comparisons across Braak stages suggested stage-associated shifts in cholesterol, complement, and oxidative phosphorylation pathways, consistent with progressive remodeling of metabolic and immune programs. These findings are consistent with AD being a regionally patterned, rather than transcriptionally uniform, disease. They provide a framework for prioritizing region-specific molecular pathways that may inform future therapeutic exploration.

## INTRODUCTION

Alzheimer’s disease (AD) remains one of the most formidable challenges in modern neuropathology, affecting tens of millions of individuals worldwide and consuming immense societal resources, yet mechanisms that initiate and propagate neuronal dysfunction remain incompletely understood [14]. Despite major efforts directed toward amyloid-β and tau pathology, converging evidence suggests that these hallmark lesions do not fully account for the clinical variability or the selective vulnerability observed across the human cortex [14]. Rather than progressing as a homogeneous pathological process, AD unfolds along anatomical, cellular, and molecular gradients, raising the possibility that distinct cortical regions exhibit differential vulnerability to Alzheimer’s disease [13,17].

Among higher-order association cortices, the insular cortex has emerged as a compelling component in AD pathology. Its broad interconnections and integral role in integrating interoception, emotional salience, and autonomic regulation place it at the convergence of systems perturbed in AD [14]. Neuroimaging studies consistently reveal atrophy and altered connectivity in the insula, yet the molecular origins of these functional disruptions remain unclear [17]. In contrast, Brodmann area 32 (BA32) of the anterior cingulate cortex participates in executive and affective control but appears less transcriptionally perturbed in AD, suggesting a fundamentally different response to pathological stimuli [17].

Modern transcriptomic technologies now provide a means to interrogate these region-specific differences at scale. However, most bulk RNA-sequencing analyses either pool samples across brain regions, thereby diluting spatial specificity; or focus on canonical AD-affected regions without a direct molecular comparison to functionally adjacent areas [17]. In parallel, machine learning approaches capable of discerning subtle, multivariate patterns in gene expression have advanced substantially, enabling biologically interpretable predictions of disease states that go beyond lists of differentially expressed genes [8,9,19,20].

The present study integrates these methodological advances to better characterize the transcriptional dysregulation in the insula, when compared to other cortical regions, such as BA32. Unlike the original publication, our analysis introduces region-specific pathway activity scoring rather than gene level comparisons using interpretable machine learning models trained on pathway features to classify AD status, and cross-regional feature transfer analyses to identify conserved versus region-specific molecular signatures. These additions provide region-resolved, pathway-level perspectives and predictive models that extend beyond what was available from prior gene-level analyses of this dataset. By combining differential gene expression [3–5], pathway activity scoring [6,7,10–12], interpretable machine learning [8,9,20], and cross-regional prediction, we delineate region-specific and shared molecular signatures that define AD at the transcriptional level. Our findings reveal a striking divergence in molecular architecture between the insula and BA32 and demonstrate that relatively concise sets of pathway perturbations are sufficient to classify AD samples and capture aspects of neuropathological severity [13,14].

## 2. METHODS

### 2.1 Study Design and Overview

Bulk RNA-sequencing data were obtained from the publicly available dataset GSE261050 in the NCBI Gene Expression Omnibus (GEO)[1,2]. This dataset comprises post-mortem cortical tissue collected from human donors spanning a spectrum of Alzheimer’s disease (AD) neuropathology and non-demented controls. Two anatomically distinct cortical regions were profiled: the dorsal anterior cingulate cortex, corresponding to Brodmann area 32 (BA32), and the insular cortex. Each sample includes annotations specifying donor identity, clinical diagnosis, brain region, sex, age at death, RNA integrity number (RIN), postmortem interval (PMI), extraction and sequencing batch variables, and neuropathological staging summarized by the B & B / Braak score[13]. Not all donors provided tissue from both regions, resulting in an unbalanced sampling structure. All analyses were performed at the individual-sample level. Donor identity was incorporated into pooled differential expression models and all predictive models to prevent information leakage when a single donor contributed more than one sample.

A schematic representation of the study design, sample distributions, and analysis modules is shown in Figure 1.

**Figure 1.**
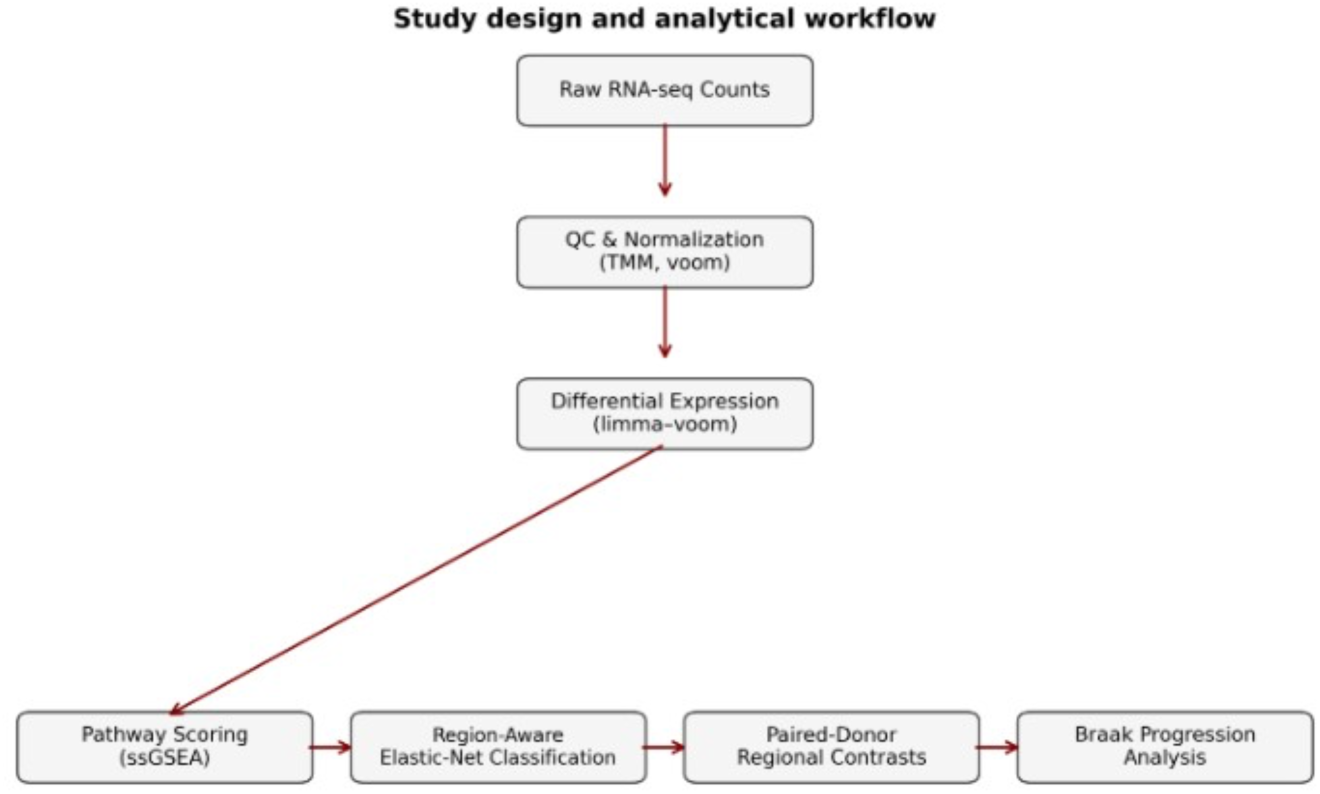
Study design and analytical workflow.

RNA-sequencing counts were processed, quality controlled, and normalized using the trimmed mean of M-values (TMM) and voom transformation [4,5]. Differential expression was then performed using limma–voom [4,5] on the normalized expression matrix. The normalized matrices were then converted into pathway activity scores using single-sample gene set enrichment analysis (ssGSEA) [6]. Pathway-level features were incorporated into region-aware elastic-net models [8] trained with donor-aware cross-validation to classify Alzheimer’s disease status. Subsequent analyses examined within-donor regional differences between BA32 and insula and evaluated pathway trajectories across Braak stages to assess the progression and regional specificity of Alzheimer’s disease pathology [13,14].

### 2.2 RNA-seq Processing and Quality Control

Pre-normalized gene-level counts and metadata were imported and harmonized prior to downstream analysis. Gene filtering was performed using edgeR’s filterByExpr function in the region-specific models [3], which removes genes lacking sufficient read support across diagnostic groups. No additional fixed count-per-million (CPM) threshold was imposed beyond this data-driven filtering step. Counts were normalized for sequencing depth using the trimmed mean of M-values (TMM) method [3], and for differential expression analyses, normalized expression values were transformed to log-counts-per-million (log-CPM) using the voom function from the limma framework [4].

Because downstream pathway scoring and visualization were performed at the gene level, Ensembl identifiers were mapped to their corresponding HGNC symbols. For these symbol-level matrices, CPM values were recomputed from TMM-normalized counts and transformed using log1p(CPM). Principal component analysis (PCA) of log-transformed expression values was used to assess the contribution of batch and other technical covariates to global variation in the data.

### 2.3 Covariate Adjustment

To mitigate the influence of technical confounders, log-transformed expression values were regressed on age, sex, RNA Integrity Number (RIN), postmortem interval (PMI), and extraction batch using linear modeling. Brain region was intentionally not included as a covariate because regional differences constitute a central biological contrast of interest. The resulting residual expression matrix was used solely for quality control evaluation. PCA performed before and after covariate regression demonstrated that this adjustment substantially reduced technical structure while preserving separation associated with diagnosis and brain region. These residuals were not used for pathway scoring or machine-learning models, which were instead derived from normalized, non-residualized expression values.

### 2.4 Differential Expression Analysis

Differential expression (DE) was performed using limma–voom [4,5] in two complementary frameworks. First, region-specific models were fit separately for BA32 and insula samples, using diagnostic group (AD vs control) as predictors. They used a donor blocked design without covariates. These models quantified transcriptional alterations within each region while adjusting for potential confounders.

Second, a pooled model leveraging both regions was fit to assess shared disease effects and region-dependent contrasts. This model included terms for diagnostic group and brain region and incorporated donor identity via the duplicateCorrelation function to account for repeated sampling of individuals [5]. Covariates were not included in the pooled donor-blocked model to keep the design focused on diagnosis and region. Potential covariate effects were evaluated separately by PCA of residualized expression values.

The pooled model provided an overall AD versus control contrast across cortex, and region-specific contrasts assessed differential responses in BA32 versus insula. Genes with false discovery rate (FDR) < 0.05 were considered significantly differentially expressed.

### 2.5 Pathway Activity Quantification

To reduce dimensionality and enhance interpretability, gene-level information was aggregated into pathway-level features using single-sample gene set enrichment analysis (ssGSEA) [7] implemented via the gseapy.ssgsea Python interface. Eight pathways with established or emerging relevance to AD were curated from prior literature that was comprised of mitochondrial oxidative phosphorylation [18], reactive oxygen species (ROS) signaling, the unfolded protein response (UPR), microglial activation [16,17], synaptic function [16], autophagy, complement activation [16], and cholesterol metabolism. For each sample, ssGSEA produced a single enrichment score per pathway, yielding an eight-dimensional profile of pathway activity.

### 2.6 Region-Aware Predictive Modeling

To test whether pathway activity could discriminate AD from controls while capturing region-dependent effects, we trained an elastic-net logistic regression classifier [8] on pathway scores. The model incorporated the eight ssGSEA pathway scores, a binary indicator specifying brain region, and interaction terms between region and each pathway score. Training and evaluation were performed using nested cross-validation with StratifiedGroupKFold[20], in which donors rather than samples were treated as groups to ensure that samples from the same individual were never split across training and test sets. Nested five-fold cross-validation was used, with five outer folds for performance estimation and five inner folds for hyperparameter tuning.

Within each outer fold, a grid search over regularization strengths (C values) was performed in inner folds to select the hyperparameter that maximized mean AUC. A final model was then fit on the full outer training set using the selected C and evaluated on the held-out test set. The primary evaluation metrics were the area under the receiver operating characteristic curve (AUROC) and average precision (AP), computed across outer folds.

### 2.7 Cross-Region Generalization

To quantify the degree to which AD-related pathway signatures were conserved across cortical areas, additional classifiers were trained exclusively on BA32 samples and evaluated on insula samples, and vice versa. Preprocessing steps, feature definitions, and hyperparameters were held constant across analyses. These experiments assessed whether the learned decision boundaries predominantly reflected global disease mechanisms or region-specific transcriptional patterns.

### 2.8 Paired-Donor Pathway Comparisons

For donors who provided tissue from both BA32 and insula, within-donor pathway differences were computed as Δ = BA32 − insula for each pathway. Because this contrast reflects biological variation independent of inter-subject differences, it provides a direct test of regional specialization. One-sample t-tests were used to determine whether Δ differed significantly from zero across donors for each pathway. To evaluate disease effects on regional differences, Welch’s t-tests were applied to compare Δ between AD and control donors.

### 2.9 Neuropathology Progression Analysis

Neuropathological severity was assessed using the B & B (Braak-like) staging variable [13] included in the original metadata. To focus on disease-related progression, analyses were restricted to AD donors. Braak scores were binned into intermediate (3–4) and advanced (5–6) pathology groups; in this dataset, no AD donors fell into the 0–2 category. For each pathway, Welch’s t-tests were used to compare ssGSEA-derived pathway enrichment scores between Braak 3–4 and 5–6 groups. These analyses provided a global assessment of stage-dependent transcriptional alterations without assuming linear relationships and reflected the available distribution of neuropathology in AD cases.

## 3. RESULTS

### 3.1 Quality Control and Covariate Adjustment

We first evaluated the extent to which technical factors contributed to variation in gene expression to determine whether covariate adjustment was necessary. Prior to covariate adjustment, PCA of log-transformed expression values showed dispersion driven primarily by non-biological variation, including extraction batch and RNA integrity number (RIN) (Figure 2A). Following regression of these technical covariates, PCA performed on residual expression values demonstrated substantially reduced technical structure while preserving biologically relevant separation associated with diagnostic status and brain region (Figure 2B). These findings confirm that covariate correction mitigated noise with minimal effect on meaningful signal and justify the use of non-residualized expression values for downstream modeling.

**Figure 2.**
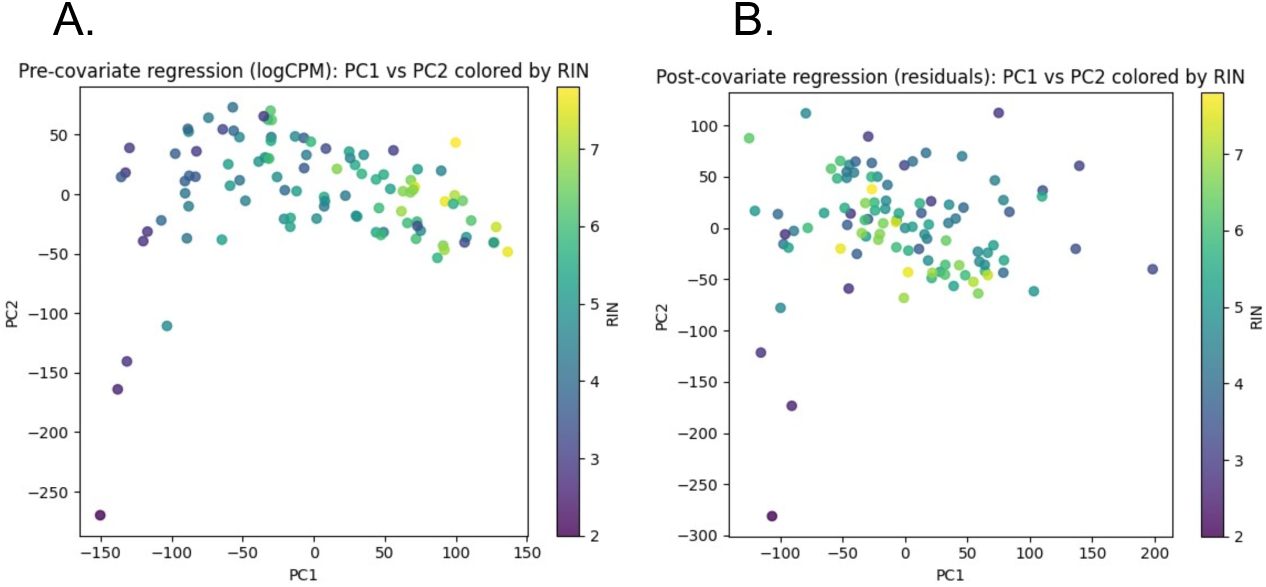
Effects of covariate adjustment on transcriptomic structure. (A) Principal component analysis (PCA) of log-transformed expression values prior to covariate regression shows clustering driven primarily by technical variables such as extraction batch and RNA integrity number. (B) PCA of residuals after regressing age, sex, RNA integrity number, postmortem interval, and batch reveals reduced technical structure and clearer separation associated with diagnostic group and brain region, supporting the use of non-residualized values for downstream modeling.

### 3.2 Differential Expression Across Regions

We next examined whether Alzheimer’s disease produced distinct transcriptional effects in different cortical regions. Region-specific models (donor-blocked, no additional covariates) identified widespread transcriptional dysregulation in the insula, with 4,883 genes significantly differentially expressed at FDR < 0.05, whereas no genes reached significance in BA32 under the same criteria. The pooled donor-blocked model confirmed a cortex-wide AD signature and further demonstrated that the magnitude of AD effects was significantly larger in insula than in BA32 (Figure 3). In this pooled model, BA32 and insula samples were analyzed together in a single donor-blocked design to assess shared AD-associated transcriptional alterations across cortical regions.

**Figure 3.**
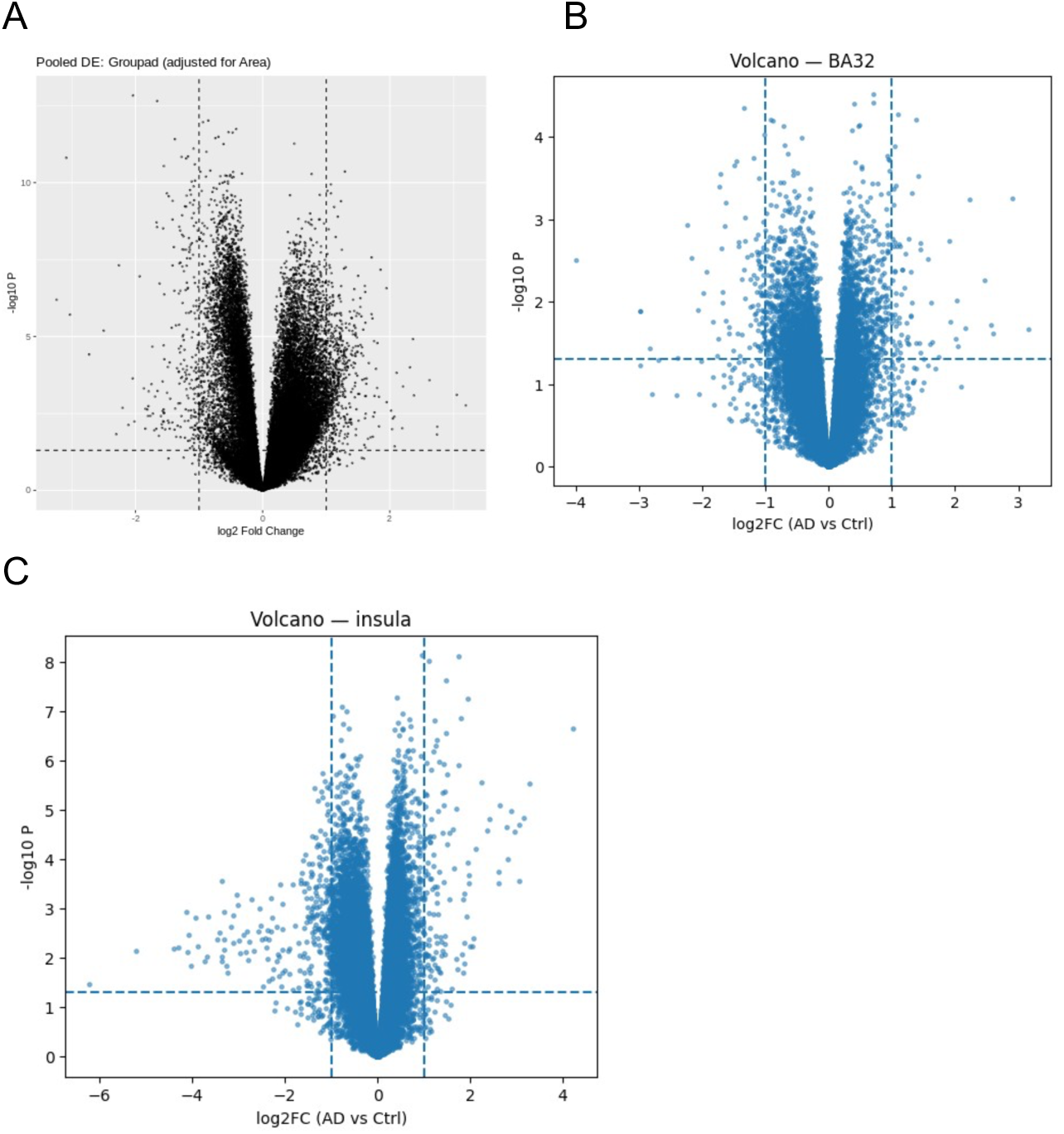
Differential expression across cortical regions in Alzheimer’s disease. (A) Pooled AD versus control contrast from a donor-blocked limma–voom model including brain region as a factor. (B) Region-specific differential expression in BA32, donor blocked models without additional covariates (C) Region-specific differential expression in insula using the same adjustment model. These complementary analyses show extensive AD-related dysregulation in insula, whereas BA32 exhibits no genes meeting FDR < 0.05.

### 3.3 Pathway Activity Differences

As a result of gene-level differences being highly region-specific, we next examined pathway activity to assess whether coordinated biological processes captured diagnostic and regional effects more robustly than individual genes. Pathway-level analyses demonstrated consistent differences associated with both diagnosis and brain region. Several pathways implicated in mitochondrial dysfunction, proteostasis, inflammation, and synaptic integrity exhibited altered activity in AD. Across all samples, microglial activation, ROS, and complement pathways tended to show higher activity in AD relative to controls, whereas oxidative phosphorylation and synaptic signaling were reduced. When stratified by region, inflammatory and complement pathways were more strongly upregulated in insula than in BA32, while synaptic and oxidative phosphorylation deficits were detectable in both regions, but again more pronounced in insula. These observations provided direct motivation for the inclusion of region-by-pathway interaction terms in predictive modeling.

**Figure 4.**
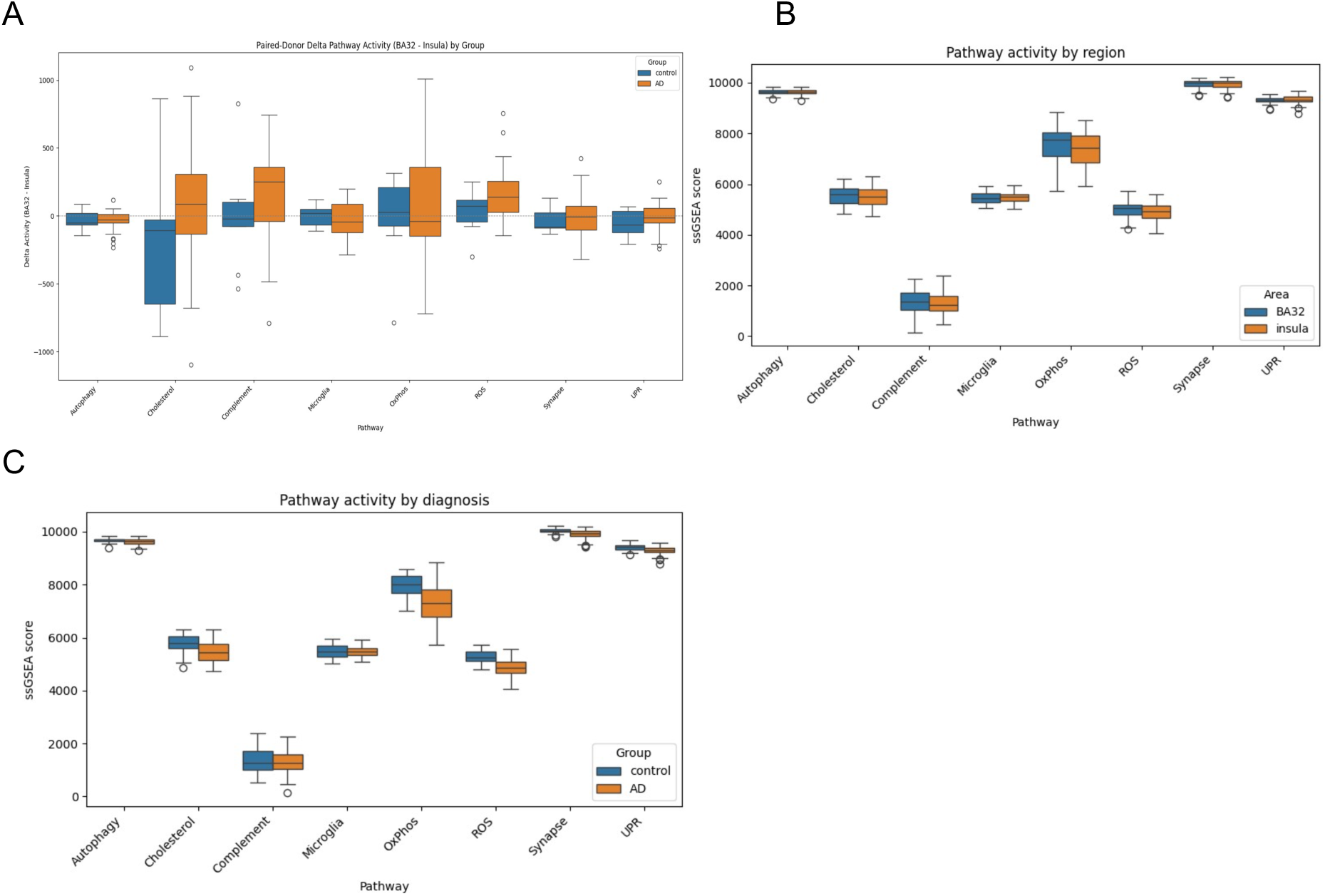
Pathway activity patterns in Alzheimer’s disease and across cortical regions. (A) ssGSEA pathway activity scores comparing AD and control samples reveal increased microglial activation, complement signaling, and reactive oxygen species pathways in AD, with reduced oxidative phosphorylation and synaptic signaling. (B) Region-stratified pathway scores demonstrate that these alterations are more pronounced in insula than BA32, particularly for inflammatory and immune-associated pathways. (C) Pairwise pathway correlation structure illustrates coordinated upregulation of immune pathways and concurrent downregulation of metabolic and synaptic pathways. Together, these analyses show that AD impacts multiple biological systems in a region-dependent manner.

### 3.4 Machine Learning Classifier Performance

We next evaluated whether the observed pathway and region effects could be harnessed for diagnostic prediction by training a region-aware elastic-net classifier on ssGSEA-derived pathway scores. The region-aware elastic-net classifier trained on pathway activity, region, and interaction features reliably discriminated AD from control samples in nested donor-aware cross-validation.

Across outer folds, the model achieved a mean AUROC of 0.813 ± 0.127 and a mean AP of 0.938 ± 0.040, indicating strong diagnostic performance despite limited sample size and substantial inter-individual variability.

These findings demonstrate that a biologically constrained, low-dimensional feature representation, comprising only eight pathways and a region term, is sufficient to recover robust AD-related signals when appropriately regularized and evaluated in a donor-aware framework.

**Figure 5.**
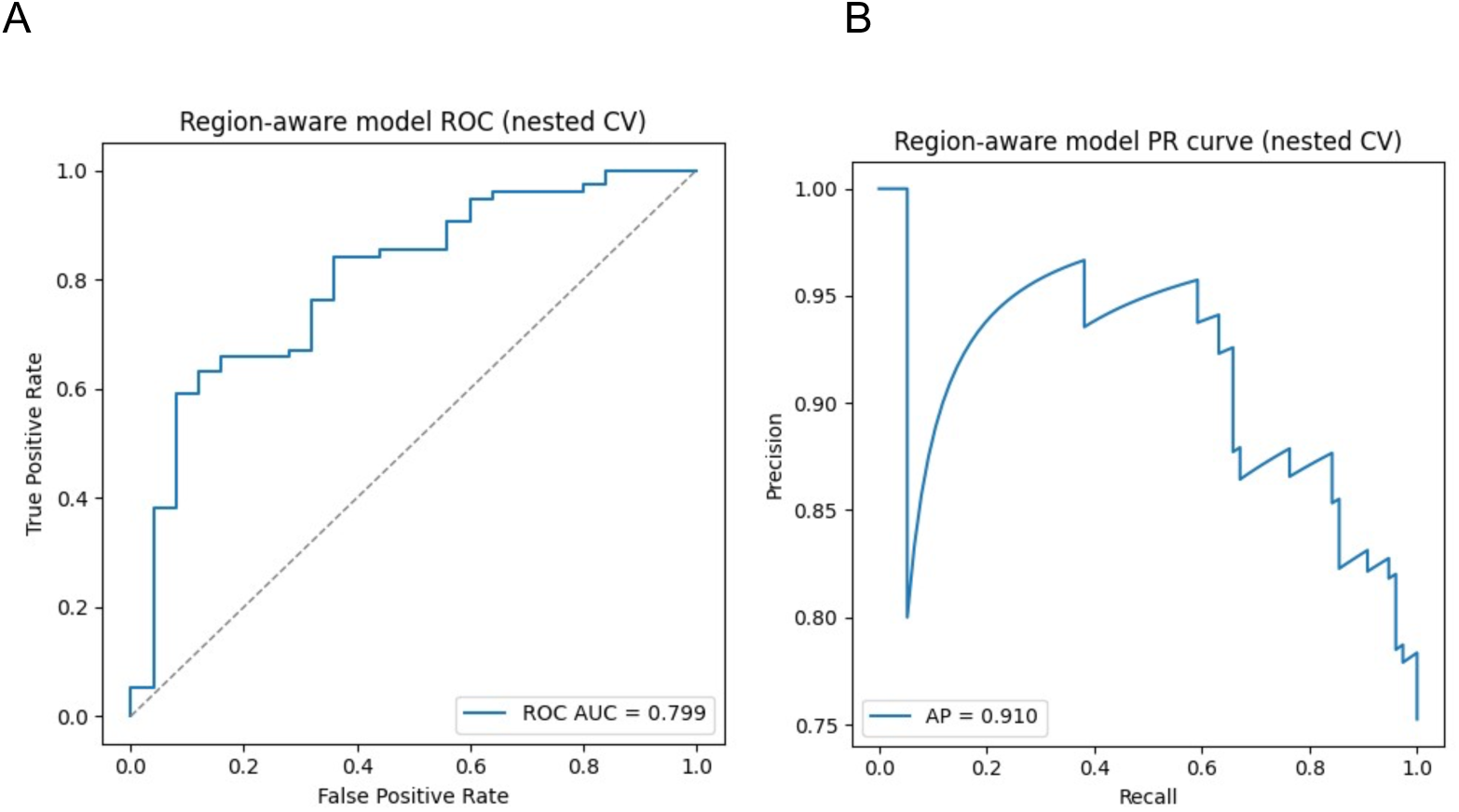
Performance of region-aware pathway-based classifiers. (A) Receiver operating characteristic (ROC) curve for the pooled region-aware model. (B) Precision–recall (PR)curve for the pooled region-aware model.. The pooled model achieved Mean AUROC = 0.813 ± 0.127 and Mean AP = 0.938 ± 0.040, indicating robust discrimination of AD versus control samples using only eight pathways and a region term.

### 3.5 Model Interpretation Using SHAP Values

To identify which pathways drove model predictions and assess their biological relevance, we interpreted the pooled classifier using SHapley Additive exPlanations (SHAP) values. Interpretation of classifier predictions using SHAP values revealed that multiple pathways exerted nonredundant contributions to the decision function. Pathways related to microglial activation, mitochondrial oxidative stress, and synaptic signaling exhibited among the strongest influences on predicted AD probability. SHAP beeswarm plots indicated that the directionality and magnitude of pathway effects varied across samples in a manner consistent with regional context, with inflammatory and complement activity contributing more strongly to AD classification in insula-enriched samples, and synaptic and oxidative phosphorylation deficits contributing across both regions.

**Figure 6.**
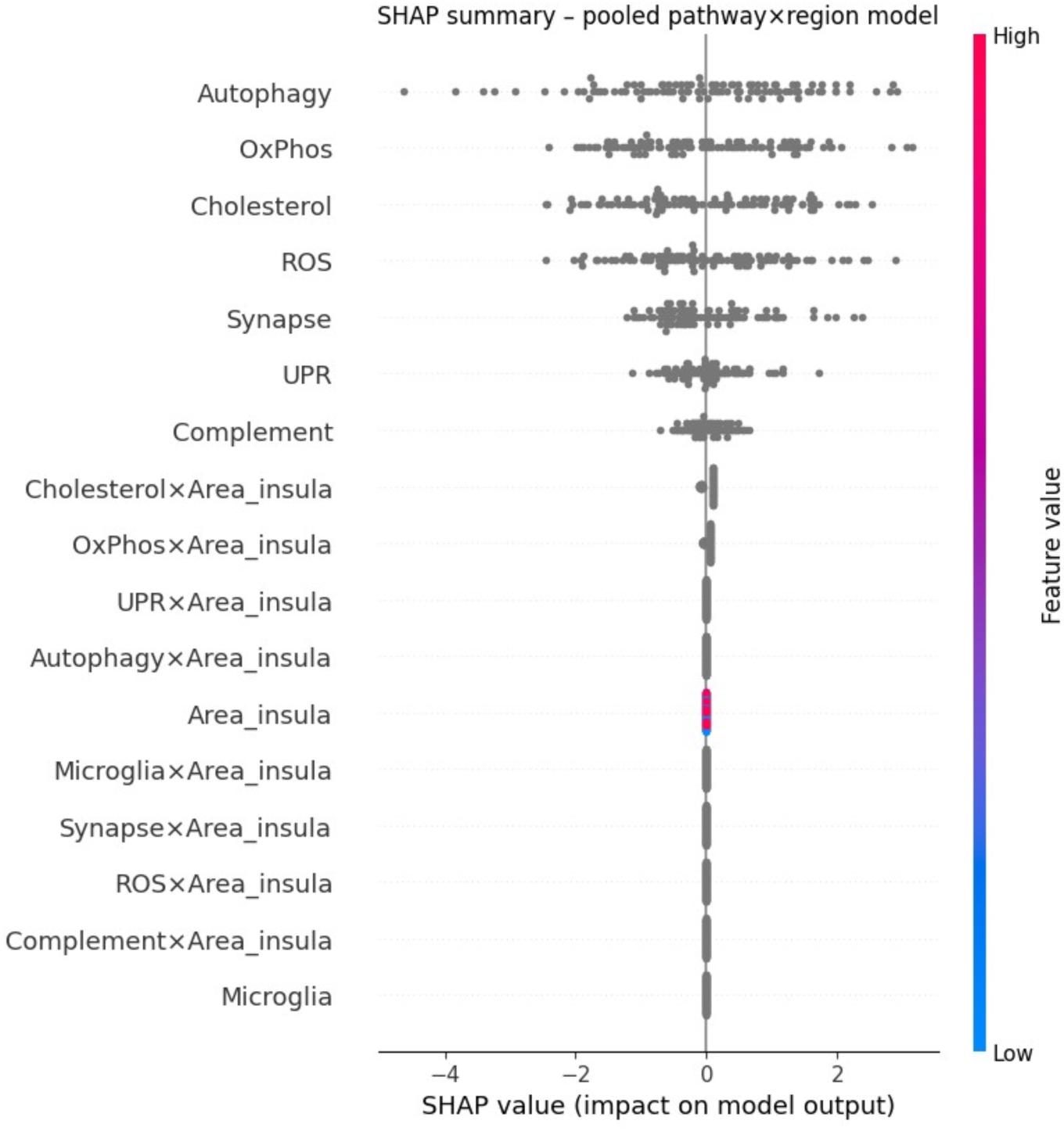
Model interpretability using SHAP values. Model interpretability using SHAP values. Mean absolute SHAP values reveal the dominant contribution and directionality of pathway features to AD classification, with inflammatory, complement, and ROS-related pathways exerting the strongest effects, particularly in insula samples. These results demonstrate that pathway level signatures are not only predictive but mechanistically interpretable.

### 3.6 Cross-Regional Generalization

To determine whether pathway-based AD signatures are region-specific or reflect shared disease mechanisms that transfer across cortical areas, we evaluated the ability of models trained in one region to predict AD status in another. Models trained in one cortical region retained discriminative performance when evaluated in the other, although accuracy declined compared with within-region evaluation. BA32-trained models applied to insula and insula-trained models applied to BA32 both performed above chance, indicating that a subset of pathway signatures generalizes across cortical areas. However, the performance decrement relative to within-region training underscores that key aspects of the AD transcriptomic signature are region-dependent and cannot be fully captured by a single, region-agnostic decision boundary.

**Figure 7.**
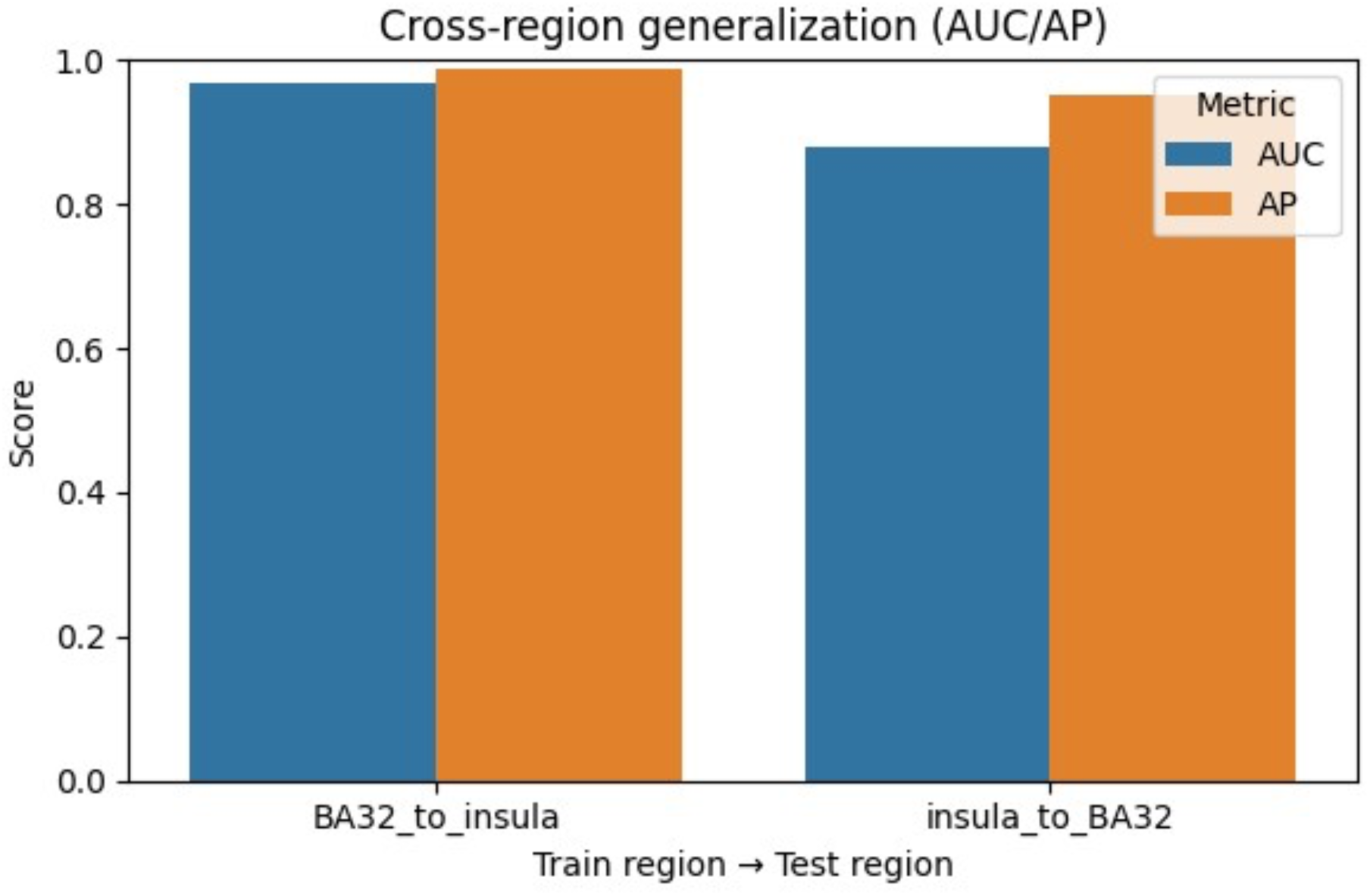
Cross-regional generalization of pathway-based models. Models trained in one region and tested in the other (BA32→insula; insula→BA32) retain above-chance classification performance, although accuracy declines relative to within-region testing. This indicates that both global and region-restricted components of the AD transcriptomic signature contribute to predictive performance.

### 3.7 Region-Specific Pathway Models

Elastic-net logistic regression models trained separately within each region using the eight pathway activity scores as features yielded distinct coefficient patterns. In BA32, classifier weights emphasized synaptic and oxidative phosphorylation pathways, consistent with more subtle yet detectable alterations in these processes in this region. In insula, coefficients for microglial activation, complement, and ROS pathways were larger, mirroring the extensive inflammatory and stress-related gene-level dysregulation observed in DE analyses. These region-specific models highlight that AD engages overlapping but not identical biological mechanisms across cortical areas.

**Figure 8.**
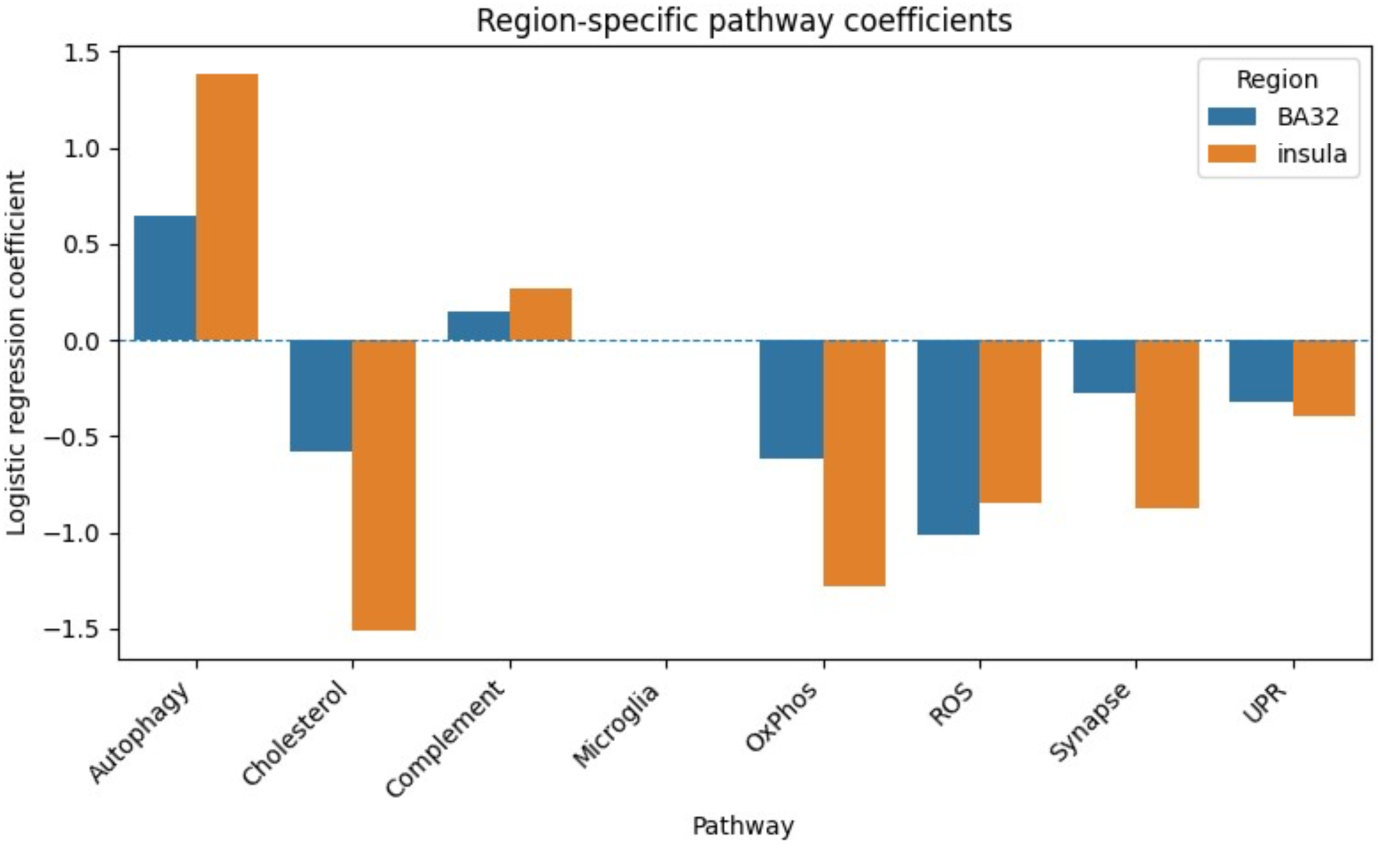
Region-specific pathway coefficient profiles. Elastic-net models trained separately in BA32 and insula yield distinct patterns of pathway importance. BA32-trained models emphasize synaptic and oxidative phosphorylation pathways, whereas insula-trained models upweight inflammatory and complement pathways, reflecting anatomically divergent disease mechanisms.

### 3.8 Paired-Donor Regional Differences

Comparison of paired BA32–insula samples demonstrated that AD alters the relative activity of several pathways across regions. Forty-one donors contributed both BA32 and insula tissue, enabling computation of within-donor Δ values for each pathway. Across all donors, the mean Δ (BA32 – insula) was substantially negative for autophagy and significantly positive for ROS, indicating lower autophagy but higher ROS activity in BA32 relative to insula at the within-subject level. In AD versus control stratified analyses, no statistically significant differences in Δ were detected, although effect sizes for several inflammatory pathways trended toward larger regional gaps in AD.

**Figure 9.**
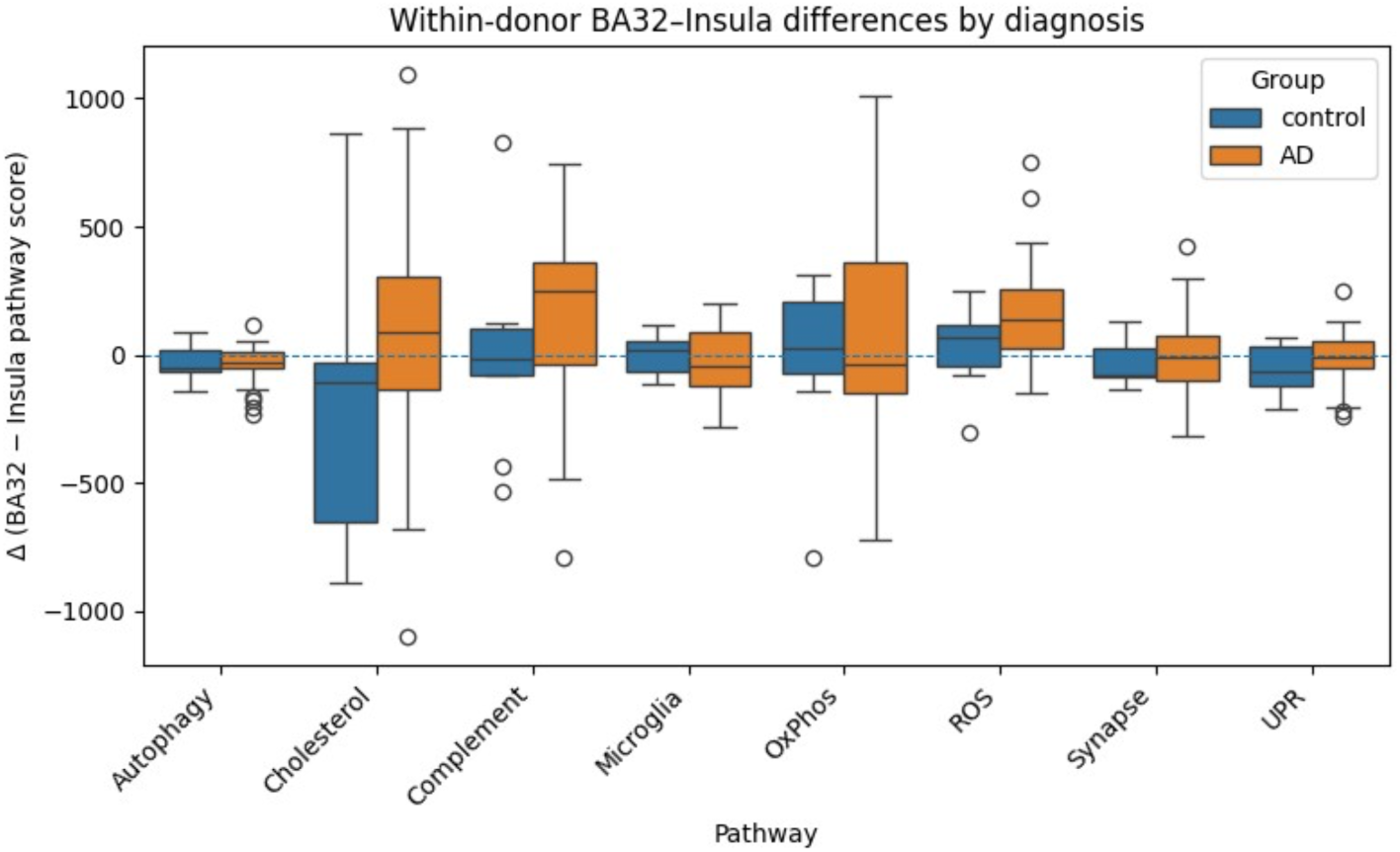
Paired-donor pathway differences between BA32 and insula. Pathway-level differences (Δ = BA32 − insula) computed within donors show that several pathways differ regionally, independent of inter-individual variability. While Δ values tended to be larger in AD donors for some inflammatory pathways, these group differences were not statistically significant, and should be interpreted as qualitative trends.

### 3.9 Pathway Activity Across Braak Stages

To evaluate the relationship between neuropathological severity and pathway activity, we examined ssGSEA scores as a function of Braak staging. Although nondemented control donors were present in the 0–2 bin, no AD donors in this dataset fell into that category; therefore, inferential analyses were restricted to AD donors falling within Braak stages 3–4 (intermediate pathology) and 5–6 (advanced pathology). Across AD donors, Welch’s t-tests revealed significantly higher activity of cholesterol and complement pathways and lower activity of oxidative phosphorylation in Braak 5–6 compared with 3–4. Microglial and synaptic pathways showed directionally consistent but nonsignificant trends, whereas ROS, autophagy, and unfolded protein response did not differ significantly between stages.

Descriptive heatmaps and boxplots (Figure 10A–B) illustrate global shifts across Braak stages, while region-stratified trajectories (Figure 10C–J) suggest that the magnitude of pathway disruption often differs between BA32 and insula. Inflammatory and immune-associated pathways appear to increase more sharply in the insula, whereas oxidative and synaptic alterations are present in both regions but visually more pronounced in the insula. Together, these patterns are consistent with stage-associated, region-sensitive remodeling, though formal tests of region-by-stage interaction are beyond the scope of this analysis. Pathway trajectory analysis revealed that complement and innate immune programs were within the Braak stages represented here, immune pathways show the steepest and earliest-detectable divergence relative to other systems, suggesting they may be among the first transcriptional programs to shift during pathological escalation, although the absence of AD donors in Braak 0–2 precludes definitive inference about disease initiation.

**Figure 10.**
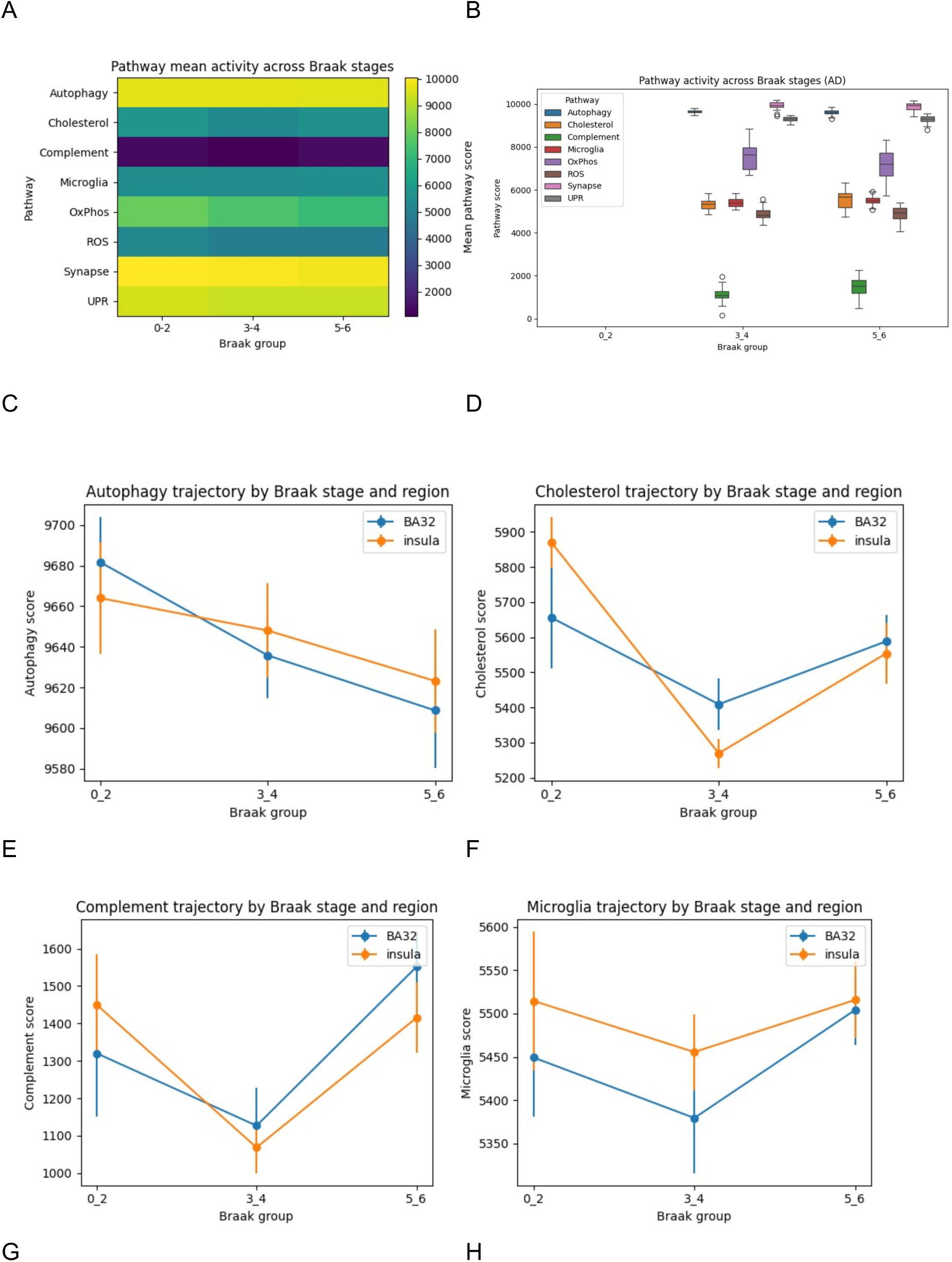
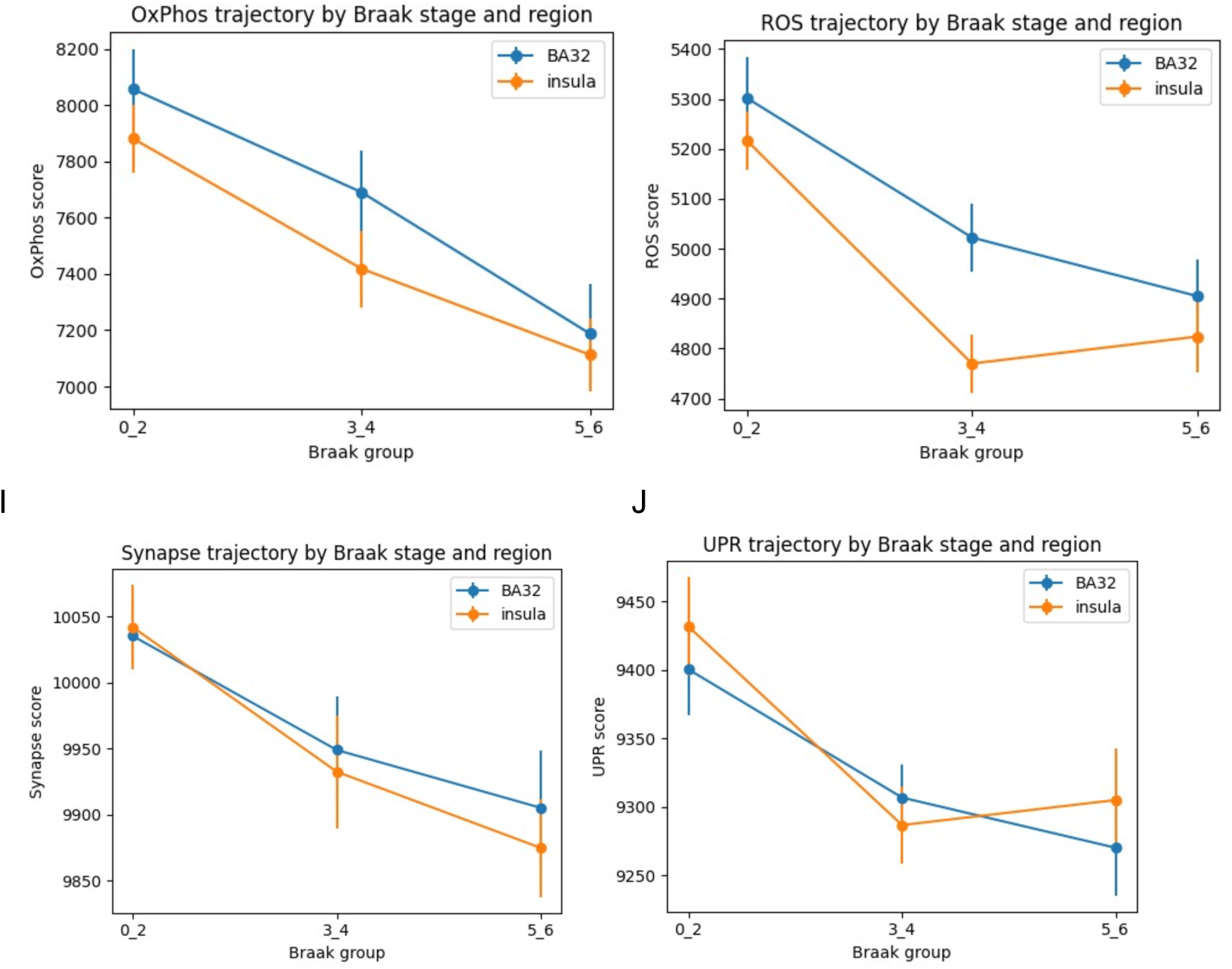
Pathway dynamics across Braak stages of Alzheimer’s disease. (A) Heatmap of mean ssGSEA pathway activity across Braak stage groups. No AD donors belonged to Braak stage 0–2; values for this bin derive from nondemented controls and are included for reference only. (B) Boxplots of pathway activity across Braak stages. (C–J) Region-stratified pathway trajectories for Autophagy, Cholesterol, Complement, Microglial activation, Oxidative phosphorylation, Reactive oxygen species, Synaptic signaling, and Unfolded protein response pathways. Inflammatory and immune pathways progressively increase with disease severity, whereas metabolic and synaptic pathways decline, with effects strongest in the insula. These results are consistent with AD pathway disruption being both progressive and anatomically compartmentalized.

## DISCUSSION

In this study, we investigated whether Alzheimer’s disease presents as a uniform cortical phenotype or whether its molecular features differ according to brain region and disease stage. We integrated differential expression analysis, pathway activity profiling, Braak-stage progression analysis, and interpretable machine learning to compare transcriptional alterations in the insula and BA32, two association cortices involved in higher-order cognitive and affective functions. Our results revealed substantial regional differences, with the insula showing extensive gene-level dysregulation and clear pathway perturbations, while BA32 displayed no significant gene-level changes, yet still exhibited modest but detectable shifts in pathway activity. We also observed that several pathways changed progressively across Braak stages, with inflammatory and metabolic pathways emerging early and increasing in magnitude with disease severity. Finally, pathway-based classifiers accurately distinguished AD samples and showed partial generalization across regions, indicating that some disease-associated signatures are shared but expressed to different degrees depending on cortical context. These findings challenge the idea of Alzheimer’s disease as a single, homogeneous molecular entity and demonstrate that both regional identity and disease stage influence its transcriptomic landscape.

The pronounced divergence in transcriptional architecture between the insula and BA32 challenges the traditional portrayal of AD as a uniformly expressed molecular entity[14]. Instead, our analyses reveal a landscape in which regional identity may dictate the scale and nature of molecular dysregulation. The insula appears more susceptible to extensive transcriptional dysregulation, while BA32 exhibits minimal gene-level dysregulation at FDR < 0.05, despite detectable pathway-level shifts.

This vulnerability likely reflects the insula’s status as a convergence hub linking internal bodily states with affective and cognitive representations [14]. High metabolic demand and structural specialization, including the presence of unique neuronal subtypes, may render the insula less tolerant of AD-associated stressors. The region’s transcriptional response encompasses pathways implicated in neuroimmune activation, metabolic failure, and synaptic dysfunction, aligning with multifactorial models of AD that extend beyond the primary drivers of amyloid or tau accumulation [14,16–18].

The ability of pathway-based models to classify AD samples underscores the diagnostic potential of interpretable, multiscale molecular signatures [6–9,20]. Importantly, the partial generalization of insular signatures to BA32 suggests that certain disease-associated perturbations may percolate across cortical networks even when not expressed at a scale detectable by gene-level tests. These observations support a model in which AD engages common molecular motifs, but their manifestation varies regionally according to local cellular architecture and functional demands [13,17].

Our findings also highlight an underappreciated opportunity: regions such as BA32, which preserve transcriptional equilibrium despite pathological exposure, may harbor intrinsic protective mechanisms. Understanding these resilience factors may prove as valuable as elucidating drivers of vulnerability and could help guide the development of neuroprotective strategies [17,18].

Together, these data indicate that Alzheimer’s disease may not arise as a uniform, cortex-wide failure of neuronal or metabolic programs [14]. Within the Braak stages represented here, the most pronounced pathway alterations are observed in the Insula, where complement and innate immune pathways become dysregulated more prominently than metabolic or synaptic alterations at comparable pathological stages [16,17], raising the possibility that immune perturbations could be positioned upstream in the detectable trajectory of cortical remodeling [16]. The fact that the dataset lacks AD donors in early Braak stages (0–2) [13], these analyses cannot determine whether immune dysregulation represents an initiating event or an early response.

## CONCLUSION

This study provides evidence that Alzheimer’s disease is not a single transcriptional phenotype but a spatially patterned molecular process whose severity and composition depend on cortical identity [14]. The insula undergoes extensive pathway-level reprogramming, whereas BA32 remains transcriptionally stable by conventional criteria. Machine learning reveals that despite this stability, BA32 contains subtle echoes of AD-associated pathway shifts detectable only in aggregate [8,9,20]. These findings define a mechanistic bridge between regional vulnerability, molecular remodeling, and disease progression, establishing a framework for identifying region-targeted therapeutic interventions and refining biomarker strategies based on anatomical context.

## Data Availability

All data produced in the present work are contained in the manuscript

## Competing Interests

BEP holds equity in Pythia Biosciences.

## Data Availability

The original data for this study is available in the Gene Expression Omnibus: https://www.ncbi.nlm.nih.gov/geo/query/acc.cgi?acc=GSE261050.

## Funding

No external funding was obtained for this study.

## Notes

### Competing Interest Statement

BEP has equity in Pythia Biosciences.

### Funding Statement

This study did not receive any funding

### Author Declarations

https://www.ncbi.nlm.nih.gov/geo/query/acc.cgi?acc=GSE261050

